# Enhanced physician performance when using an artificial intelligence model to detect ischemic stroke on computed tomography

**DOI:** 10.1101/2023.01.16.23284632

**Authors:** James M Hillis, Bernardo C Bizzo, Romane Gauriau, Christopher P Bridge, John K Chin, Buthaina Hakamy, Sarah Mercaldo, John Conklin, Sayon Dutta, William A Mehan, Robert W Regenhardt, Ajay Singh, Aneesh B Singhal, Jonathan D Sonis, Marc D Succi, Tianhao Zhang, Bin Xing, John F Kalafut, Keith J Dreyer, Michael H Lev, R Gilberto González

## Abstract

Acute ischemic stroke can be subtle to detect on non-contrast computed tomography imaging. We show that a novel artificial intelligence model significantly improves the performance of physicians, including ED physicians, neurologists and radiologists, in identifying and quantifying the volume of acute ischemic stroke lesions. This model may lead to improved clinical decision-making for stroke patients.

## Article

The early imaging features of acute ischemic stroke on non-contrast computed tomography (CT) can be subtle. We previously reported the development of a deep learning model that detects ischemic core on CT studies and was superior to three experienced neuroradiologists.^1^ As part of its training, the model utilized segmentations obtained from the region of acute infarct on paired magnetic resonance imaging (MRI) studies that were then registered onto the CT studies. This design had recognized that MRI better detects early acute ischemic stroke but CT is a cheaper, quicker and more widely available imaging modality.

In further evaluating this model, we wished to see how its use impacted physicians who were interpreting non-contrast CT studies. We therefore designed a multi-reader multi-case study whereby physicians interpreted 180 CT cases both with and without the use of the model (Supplementary Figure 1). There were 8 physicians (2 emergency physicians, 2 emergency radiologists, 2 neurologists, 2 neuroradiologists) who each interpreted 90 cases with the model output and 90 cases without the model output (Figure 1A). After a four-week washout period, they interpreted the cases in the opposite manner (i.e., the cases that they had previously interpreted without the model output were now interpreted with the model output).

**Figure 1:**
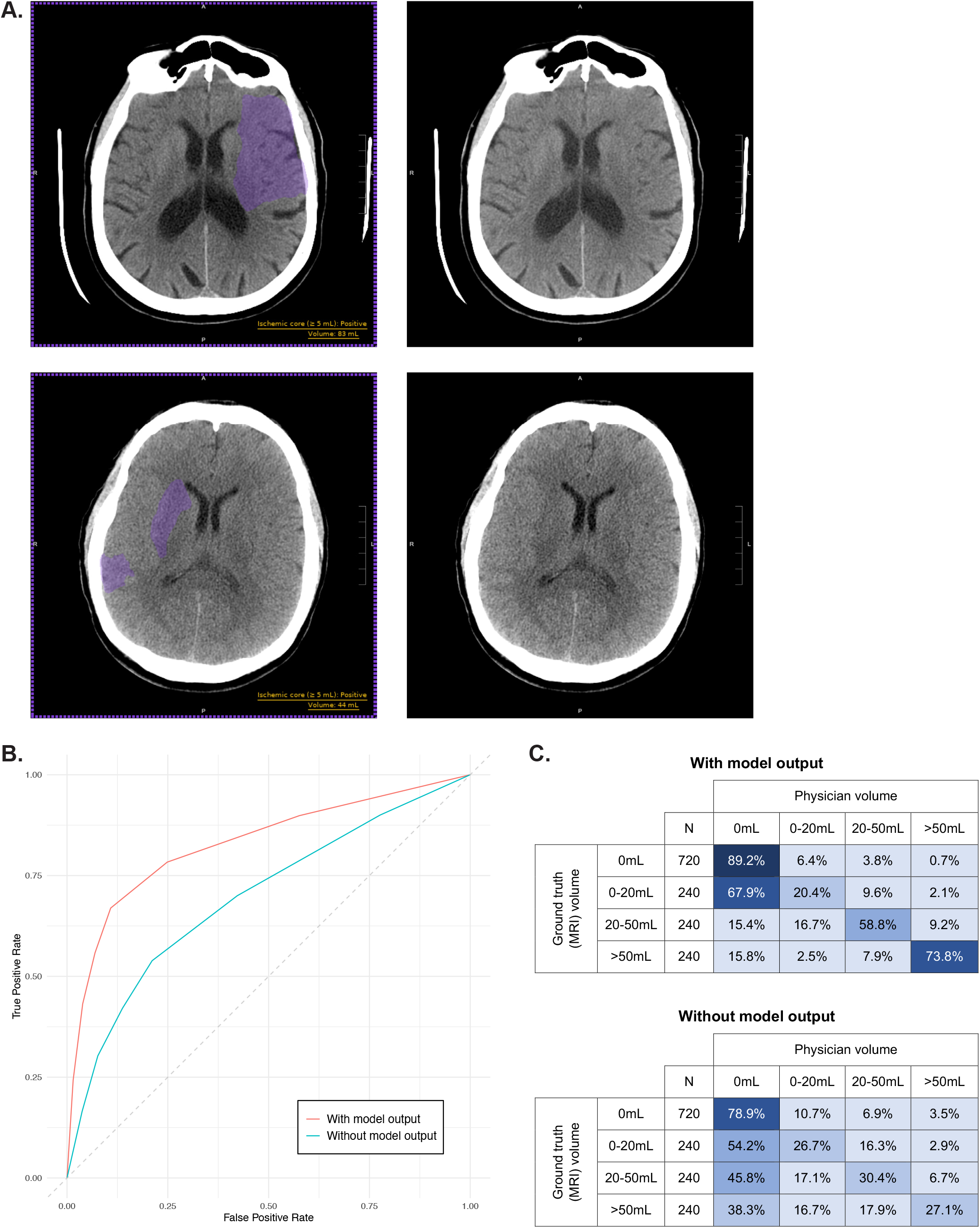
**A**. Example of two cases with and without model output showing text incorporating binary classification for ischemic core ≥5mL and volume, and purple segmented region. The first case, with infarct volume 77mL on MRI, was detected by 1 out of the 8 physicians without model output and 6 with model output. The second case, with infarct volume 45mL on MRI, was detected by 2 without model output and 8 with model output. B. Receiver operating characteristic curves for both with model output and without model output. C. Confusion matrices comparing physician volume with ground truth volume; a correct volume estimate occurs on the diagonal; a volume of 0mL reflects a negative case.

The model output included the binary classification of whether ischemic core ≥5mL was present. If there was ischemic core ≥5mL, the output also included the volume of the ischemic core and the segmented region of the ischemic core. The physicians were asked up to three multiple choice questions: the presence of any ischemic core (positive, negative), their level of certainty for the presence or absence (scale of 1-7; see Supplementary Table 1 for details), and, if they stated the presence of ischemic core, their volume estimate (0-20mL, 20-50mL, >50mL; upper bound of range considered inclusive). Their responses were compared to the ground truth interpretations demonstrated by diffusion imaging on a paired MRI case.

The physicians performed significantly better at binary detection when using the model compared with not using the model. Their area under the receiver operating characteristic curve (AUC) improved from 0.696 to 0.836 (difference 0.141; 95% CI: 0.081-0.200; p<0.001; Figure 1B and Table 1). Their sensitivity improved from 53.9% to 66.9% (difference 13.1%; 95% CI: 6.5-19.6%; p<0.001). Their specificity improved from 78.9% to 89.2% (difference 10.3%; 95% CI: 6.6-14.0%; p<0.001). Their interpretation time also improved from 62.30 seconds to 43.92 seconds (difference 18.38 seconds; 95% CI: 15.59-21.18 seconds; p<0.001).

**Table 1:**
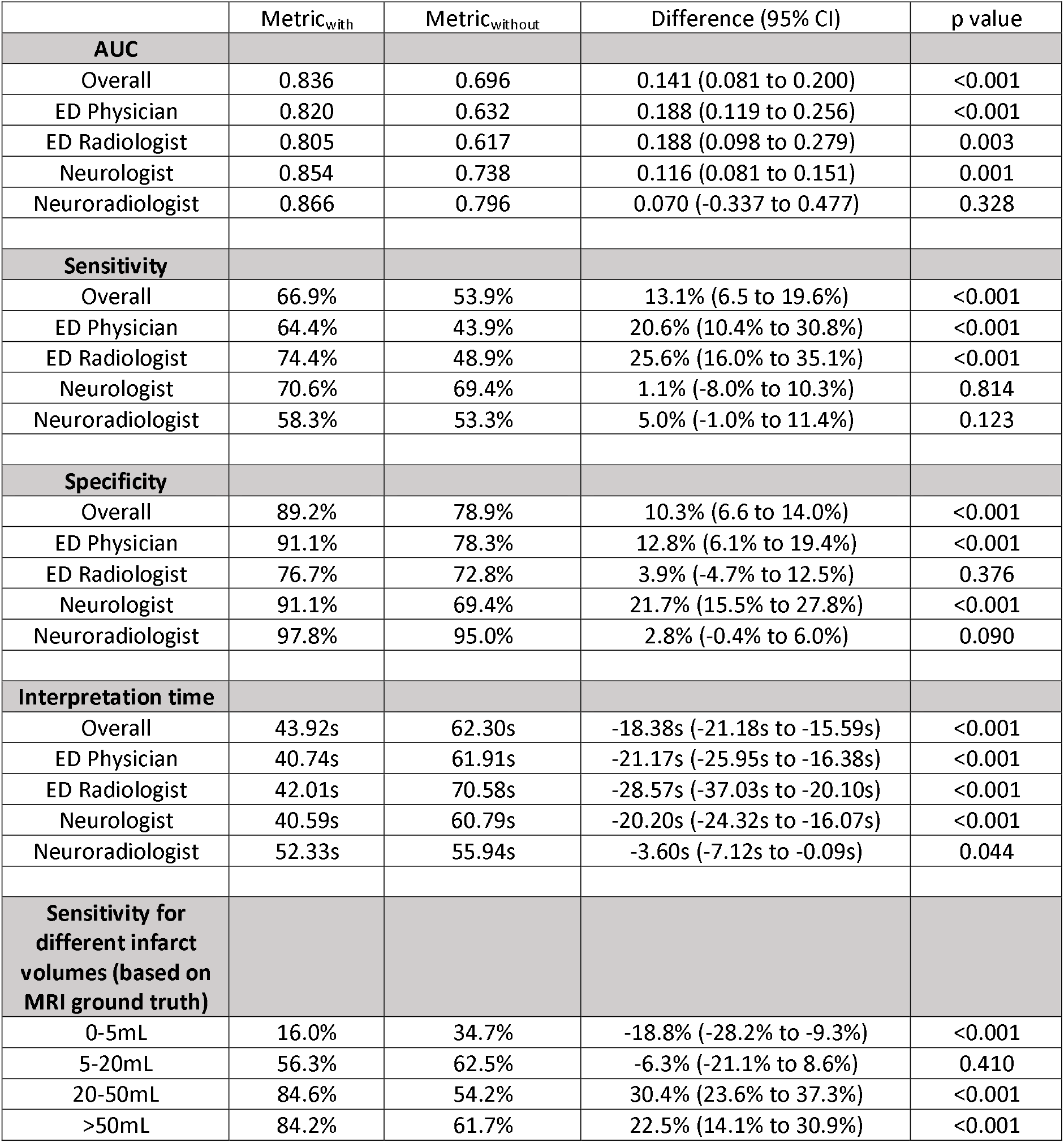
Results summary for physician performance comparing interpretations performed with or without the model outputs.

|  | Metric<sub>with</sub> | Metric<sub>without</sub> | Difference (95% CI) | p value |
| --- | --- | --- | --- | --- |
| **AUC** |  |  |  |  |
| Overall | 0.836 | 0.696 | 0.141 (0.081 to 0.200) | <0.001 |
| ED Physician | 0.820 | 0.632 | 0.188 (0.119 to 0.256) | <0.001 |
| ED Radiologist | 0.805 | 0.617 | 0.188 (0.098 to 0.279) | 0.003 |
| Neurologist | 0.854 | 0.738 | 0.116 (0.081 to 0.151) | 0.001 |
| Neuroradiologist | 0.866 | 0.796 | 0.070 (-0.337 to 0.477) | 0.328 |
| **Sensitivity** |  |  |  |  |
| Overall | 66.9% | 53.9% | 13.1% (6.5 to 19.6%) | <0.001 |
| ED Physician | 64.4% | 43.9% | 20.6% (10.4% to 30.8%) | <0.001 |
| ED Radiologist | 74.4% | 48.9% | 25.6% (16.0% to 35.1%) | <0.001 |
| Neurologist | 70.6% | 69.4% | 1.1% (-8.0% to 10.3%) | 0.814 |
| Neuroradiologist | 58.3% | 53.3% | 5.0% (-1.0% to 11.4%) | 0.123 |
| **Specificity** |  |  |  |  |
| Overall | 89.2% | 78.9% | 10.3% (6.6 to 14.0%) | <0.001 |
| ED Physician | 91.1% | 78.3% | 12.8% (6.1% to 19.4%) | <0.001 |
| ED Radiologist | 76.7% | 72.8% | 3.9% (-4.7% to 12.5%) | 0.376 |
| Neurologist | 91.1% | 69.4% | 21.7% (15.5% to 27.8%) | <0.001 |
| Neuroradiologist | 97.8% | 95.0% | 2.8% (-0.4% to 6.0%) | 0.090 |
| **Interpretation time** |  |  |  |  |
| Overall | 43.92s | 62.30s | -18.38s (-21.18s to -15.59s) | <0.001 |
| ED Physician | 40.74s | 61.91s | -21.17s (-25.95s to -16.38s) | <0.001 |
| ED Radiologist | 42.01s | 70.58s | -28.57s (-37.03s to -20.10s) | <0.001 |
| Neurologist | 40.59s | 60.79s | -20.20s (-24.32s to -16.07s) | <0.001 |
| Neuroradiologist | 52.33s | 55.94s | -3.60s (-7.12s to -0.09s) | 0.044 |
| **Sensitivity for different infarct volumes (based on MRI ground truth)** |  |  |  |  |
| 0-5mL | 16.0% | 34.7% | -18.8% (-28.2% to -9.3%) | <0.001 |
| 5-20mL | 56.3% | 62.5% | -6.3% (-21.1% to 8.6%) | 0.410 |
| 20-50mL | 84.6% | 54.2% | 30.4% (23.6% to 37.3%) | <0.001 |
| >50mL | 84.2% | 61.7% | 22.5% (14.1% to 30.9%) | <0.001 |

These improvements were maintained across most specialties as part of a subgroup analysis (Table 1). The physicians with the greatest improvement were emergency physicians and emergency radiologists, which is consistent with them having the least experience in interpreting brain imaging and therefore having the greatest opportunity to benefit. The physicians with the least improvement were neuroradiologists, who have the most experience.

The improvement in sensitivity was most pronounced for larger infarct volumes (Table 1). The physicians’ sensitivity improved from 54.2% without the model output to 84.6% with the model output for 20-50mL infarcts, and 61.7% to 84.2% for >50mL infarcts. These large infarcts are more likely to involve a large vessel occlusion, which means the patients could benefit from treatment with endovascular thrombectomy.^2-8^ The increased ability to detect these infarcts could prompt physicians to obtain CT angiography to detect a large vessel occlusion and cue the physicians interpreting the CT angiography to the likely vessel involved. This increased ability to detect may also lead to improved triage and sooner evaluation for endovascular thrombectomy. A decreased time to thrombectomy has previously been shown to improve outcomes.^9^

There was, however, a decline in sensitivity for infarct volumes 0-5mL (Table 1). This decreased performance was expected given that the model only outputs whether it has detected ischemic core ≥5mL (i.e., so the model should classify these cases as negative). The reason for the model using this volume threshold is to avoid false positive interpretations from small regions of noise on a CT study; the clinical concern is for such interpretations to increase MRI utilization to evaluate for infarct more definitively. While we acknowledge the subsequent decreased sensitivity amongst physicians for these infarct volumes, it is important to recognize that this study occurred outside of the clinical environment where other factors, especially the acute onset of neurologic symptoms, could alert physicians to the occurrence of ischemic stroke.

The physicians also performed better at volume quantification when using the model (Figure 1C). They correctly identified the volume range 73.8% of the time for >50mL infarcts with the model output compared to 27.1% without the model output. They correctly identified the volume range 58.8% of the time for 20-50mL infarcts with the model output compared to 30.4% without the model output.

A key consideration moving forward is how the model might impact clinical workflow. Currently the clinical paradigm is that an ischemic stroke should be assumed when a patient presents with stroke-like symptoms and a non-contrast CT does not reveal an abnormality; the key reason for obtaining the CT is to exclude intracranial hemorrhage. This study demonstrates the model improves physicians’ detection of ischemic core. Further research should be conducted to determine whether the CT could be used more than it currently is for confirmation of ischemic core. For instance, confirmation of ischemic core on non-contrast CT could be particularly helpful when a patient’s symptoms are ambiguous and ischemic stroke has not been considered as a likely differential diagnosis. The presence of ischemic core could help trigger and triage the next management steps such as CT angiography, administration of thrombolytic medication and consideration of endovascular thrombectomy.^10^

The model most helped physicians with less experience in interpreting brain imaging and may be most beneficial in rural areas with fewer subspecialty physicians. The model may separately be most helpful at hospitals that can perform non-contrast CT but do not have emergent access to advanced imaging modalities like CT perfusion or MRI. It may therefore assist in reducing the urban-rural inequities in acute stroke care.^11^

A limitation of this study was that the cases were taken from the test set from model development. While these cases were sequestered and not exposed to the model during development, they are likely to be the most similar to the training cases and therefore provide the best model performance. The assessment of generalizability of the model will benefit from evaluation on a more diverse dataset prior to clinical use. We note that the physicians were not aware of the standalone model performance on these cases during this study.

Overall, this study demonstrates how the use of an artificial intelligence model enhances physicians’ identification and volume quantification of ischemic core on non-contrast CT. It suggests that the model could provide benefit in the acute stroke clinical environment.

## Methods

### Study design

This retrospective multi-reader multi-case study was conducted using radiology cases from hospitals within the Mass General Brigham network. It was approved by the Mass General Brigham Institutional Review Board with waiver of informed consent per the Common Rule. It was conducted in accordance with relevant guidelines and regulations including the Health Insurance Portability and Accountability Act (HIPAA). This report followed the Standards for Reporting Diagnostic Accuracy (STARD 2015) reporting guideline.

### Case selection and model inference

The 180 cases were selected from the test set that had been used at the time of model development.^1^ As described previously, they had accompanying MRI studies that were used to establish the ground truth interpretations (within 3 hours of the CT for positive cases and 5 days for negative cases). They were selected using stratified randomization from the entire primary test set such that there were 30 cases with ischemic core 0-20mL, 30 cases with ischemic core 20-50mL, 30 cases with ischemic core >50mL and 90 cases without any ischemic core. This randomization ensured there were cases on which the model performed accurately and inaccurately (see Supplementary Table 2 for standalone model performance on these cases).

For the studies without the model output, the physicians were provided with only the axial 5mm series. For the studies with the model output, the physicians were provided with the axial 5mm series and an identical series with the model output incorporated into the imaging pixel data (Figure 1A). The physicians were able to visualize these two series simultaneously in adjacent window panes. The outputs included the binary classification of whether ischemic core ≥5mL was present, and, if it was present, the volume of the ischemic core and the segmented region of the ischemic core. The volume threshold of 5mL is proposed for future clinical use given it optimizes specificity by avoiding false positive interpretations through incorrect interpretation of small regions of noise on CT.

### Physicians

The physicians were chosen to ensure representation of likely future clinical users including emergency physicians, emergency radiologists, neurologists and neuroradiologists. They were all board-certified for their relevant specialty. They were trained on the annotation tasks and completed twelve training cases. They had not been involved with model development and were not aware of prior model performance results. They were informed that specificity was prioritized over sensitivity as part of the model design given that future clinical users should similarly be aware of this fact.

### Reader study process

The multi-reader multi-case study design involved the physicians interpreting all radiology studies twice: both with and without the model output (Supplementary Figure 1). The interpretations were performed as part of two sessions that were separated by at least four weeks as recommended by the US Food and Drug Administration^12^; each study was interpreted once in each session. Within each session, half of the studies were interpreted with the model output and half of the studies without the model output; the studies were interpreted in the opposite manner for the other session. The studies were split into two batches to facilitate these interpretations. The order of studies within each session was randomized and differed for each physician. The interpretation of the first batch of studies with or without model output for each physician was also randomized.

The physicians were assisted in working through the cases by an internal web-based annotation system that required them to interpret the cases in the defined order. This annotation system incorporated the FDA-cleared eUnity image visualization software (Version 6 or higher). The physicians were firstly asked about the presence of any ischemic core (positive, negative). They were then asked their level of certainty for this ischemic core on a scale of 1-7 (see Supplementary Table 1 for options). If they stated that ischemic core was present, they were also asked to estimate the volume (0-20mL, 20-50mL, >50mL; the upper bound of these ranges was considered inclusive). The annotation system also recorded the start and stop times for the interpretation of each case. When submitting the annotations for each case, the physicians could opt to move to the next case or exit; they did not need to interpret all cases in a single sitting.

### Statistical techniques

This study was a pilot multi-reader multi-case study for this model. The predefined primary assessment was comparison of the AUC with and without the model outputs. The predefined secondary assessments were comparison of sensitivity, specificity and interpretation time with and without the model outputs. The specialty subgroup analysis, infarct volume subgroup analysis and volume quantification analysis were calculated as exploratory assessments. Given the pilot nature of this study, there was not an estimated effect size and powering was not performed. There were no missing data with the exception of the excluded interpretation times as described below.

The selection of cases involved multiple randomization procedures including for the cases selected in each batch, the interpretation order for each physician, and whether the first batch for a physician was with or without model output. This randomization used the Latin squares methodology.

The analysis of the AUC was based on the 7-point scale assessments from each physician and the ground truth interpretations based on the MRI. The overall comparison between the two methods (with and without model outputs) was derived from the difference between the mean AUCs^13^ via analysis of variance (ANOVA), taking into account the variability components: methods, physicians, cases and their interactions. The analysis was implemented by using the OR-DBM MRMC software package version 2.5 or higher.^14,15^ The ANOVA model treated both physicians and cases as random samples, to be able to draw inferences for the whole population of physicians and cases. Two-sided p-values were based on 95% confidence intervals of the difference in AUCs.

The analysis of sensitivity and specificity used the binary interpretations from each physician and the ground truth interpretations based on the MRI. The comparison between the two methods (with and without model outputs) was based on the generalized estimating equations (GEE) model^16^ by using SAS procedure PROC GENMOD, taking into account repeated observations from the MRMC design. Based on SAS GENMOD, case was treated as the repeated subject, accounting for correlations of physician and method within case, by using an exchangeable covariance structure. Two-sided p-values were based on 95% confidence intervals of the difference in sensitivity and specificity.

The analysis of interpretation time was performed in SAS using a repeated measures ANOVA. Two-sided p-values were based on 95% confidence intervals of the difference in interpretation time. Thirteen interpretation times (from 2880 interpretations) were excluded; the paired study for the same physician (i.e., the equivalent case with or without model output) was also excluded to ensure balance of such exclusions. These exclusions occurred for two reasons. Firstly, the physicians could notify study management if an interpretation time should be excluded (e.g., they were interrupted). Secondly, the annotation system initially recorded the start time for each case incorrectly; to ensure consistency across the entire cohort, the interpretation time was rederived by calculating the difference in start time with the subsequent case or the difference in stop time with the previous case (the minimum of these times was taken); the interpretation times were excluded if this derived number did not appear consistent with an expected duration (e.g., it appeared a physician only interpreted one case in a sitting so the difference in times with the previous and subsequent cases would be incorrect).

The analysis of volume outputs used the volume estimates from each physician and the ground truth interpretations based on the MRI. The frequencies of how the volume estimates matched with ranges of ground truth volumes of the two methods (with and without model outputs) were calculated using Microsoft Excel.

## Supporting information

STARD Checklist

## Data Availability

The data generated for this study contains protected patient information. Some data may be available for research purposes from the corresponding author upon reasonable request.

## Acknowledgements

The authors thank the broader Mass General Brigham Data Science Office and GE Healthcare teams for their assistance with this project.

## Data availability

The data used were obtained from hospitals within the Mass General Brigham network. Data use was approved by relevant Institutional Review Board. The data are not publicly available and restrictions apply to their use.

## Author contributions

JMH, BCB, JKC, TZ, JFK, KJD, MHL and RGG conceptualized the study. JMH, BCB, RG, CPB, JKC and BH conducted the study including producing the model output. JC, SD, WAH, RWR, AS, ABS, JDS and MDS were physician readers. SM, TZ and BX performed the randomization and statistical analysis. JMH drafted the manuscript text and figures. All authors reviewed the manuscript.

