## Supplementary Material for "Enhanced physician performance when using an artificial intelligence model to detect ischemic stroke on computed tomography"

**Supplementary Tables**

**Supplementary Table 1:** The options provided to physicians for their level of certainty about the presence or absence of ischemic core. Options 1-3 were provided if their ischemic core interpretation was “negative”. Options 4-7 were provided if their ischemic core interpretation was “positive”.

1 Completely certain that no finding is present.

2 Reasonably certain that no finding is present.

3 Somewhat certain that no finding is present.

4 Equivocal that finding is present but would proceed forward as if finding present (e.g., if radiologist, would alert clinical team).

5 Somewhat certain that finding is present.

6 Reasonably certain that finding is present.

7 Completely certain that finding is present.

**Supplementary Table 2:** The standalone model performance for the 180 cases including 90 positive and 90 negative cases.

|  |  | **Ground truth** | |
| --- | --- | --- | --- |
|  |  | Positive | Negative |
| **Model inference** | Positive | 61 | 2 |
|  | Negative | 29 | 88 |
|  |  | Sensitivity: 67.8% | Specificity: 97.8% |

**Supplementary Figure**

**Supplementary Figure 1:** Structure of multi-reader multi-case study design for four of the eight physicians. It reflects that the physicians each interpreted each case twice (e.g., cases in Batch A both with and without model output) and that they each interpreted the cases in different orders.

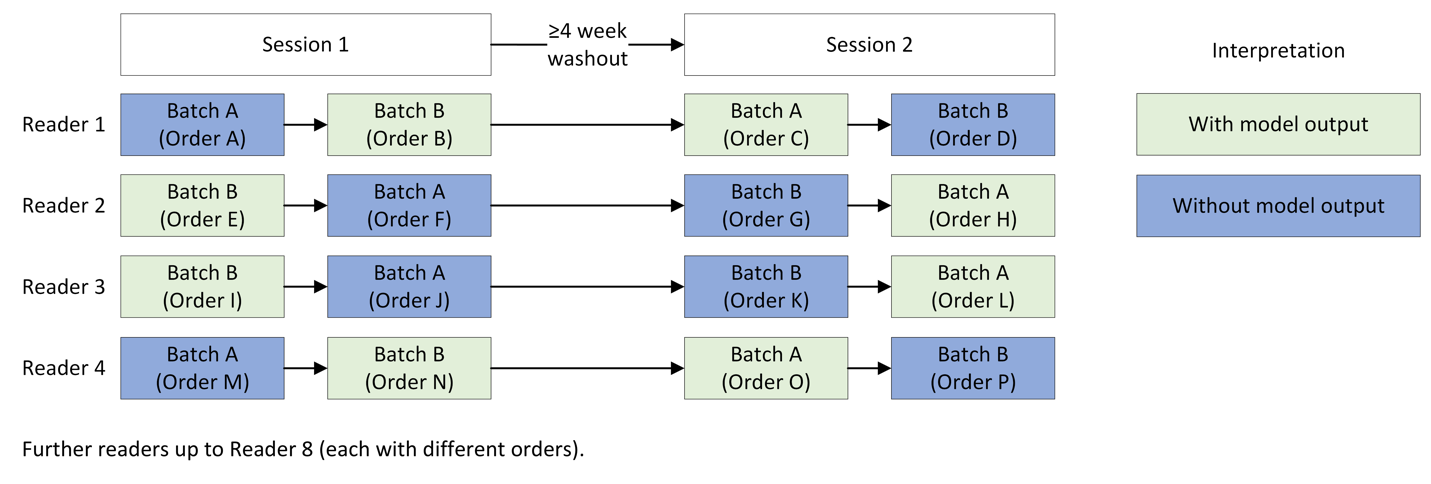
